# Intratympanic cortisone injection: an evaluation of the current data situation

**DOI:** 10.1101/2024.12.05.24318568

**Authors:** Sandra Schmidt, Milena Thomsen, Kai Johannes Lorenz

**Affiliations:** Clinic for Ear, Nose and Throat Medicine, Head and Neck Surgery and Communication Disorders at the Bundeswehr Central Hospital Koblenz

## Abstract

**Introduction:** Intratympanic cortisone injections (ITI) have been established for sudden deafness for years. ITI can be used as primary therapy and, above all, as rescue therapy if systemic administration has not brought any improvement or if there are contraindications to systemic administration.

**Methods:** Many different forms of application, indication, duration and treatment have been used. Different local anesthetics, cortisone preparations, body and head positions are practiced.

**Results:** 103 patients are analyzed. In 55 cases, the injection after oral therapy. In 48 cases, given immediately due to extreme hearing loss or comorbidities the injections are well tolerated with rare unpleasant side effects.

**Discussion:** High-dose treatment with cortisone has been used for decades. The HODOKORT study brought significant dose reductions. The KORTEBO study could not be carried out, but there is need for it. There is a lack of evidence of an optimal indication, implementation and objective proof, which is urgently needed.

## 1. Introduction

The most common indication for an intratympanic cortisone injection (ITI) is sudden deafness. Hearing loss is often defined as a hearing impairment of at least 30 dB, which is evident in the audiogram in three consecutive frequencies (1). However, this definition is not universally used as an inclusion criterion in studies because the reasons for setting the threshold are not obvious and the frequency range is not defined more precisely (2). The aetiology of sudden onset, usually unilateral sensorineural hearing loss is unknown. Viral infections, intracochlear membrane ruptures, vascular diseases, circulatory disorders and autoimmune reactions have been discussed (4). As a result, sudden hearing loss is a diagnosis of exclusion once definable causes have been diagnostically ruled out. The hearing loss may be accompanied by ringing in the ears, symptoms of dizziness or a feeling of pressure in the ear (1). In principle, sudden hearing loss can persist at any age, although the incidence increases significantly with age and is rare in childhood (3). Systemically active glucocorticoids have been used as primary therapy for 50 years, even though the benefits and therapeutic mode of action are still unclear (2). The current S1 guideline for sudden hearing loss recommends high-dose oral cortisone for 3 days with 250 mg prednisolone (5). However, the recently published multicentre HODOKORT study showed no disadvantage of significantly lower-dose cortisone administration with a lower spectrum of side effects in contrast to high-dose therapy (6). ITI glucocorticoid administration is available as a further treatment option for patients in whom systemic glucocorticoid intake has not led to sufficient regression or for whom cortisone intake is contraindicated due to pre-existing conditions. In view of the possible complications of ITI application, such as perforation of the eardrum, dizziness or pain, the question remains as to how great the benefit is for patients as the sole primary treatment option or as a combination therapy with oral glucocorticoids. Based on 103 patients from our clinic, this question will be explained and discussed in relation to previously published studies. A review of the literature reveals a completely inconsistent approach to the administration of ITI corticosteroids. A summary of the currently still inconsistent therapeutic measures will be drawn up.

## 2. Material and Methods

### 2.1. Study design

In the monocentric retrospective clinical study, 103 patients, including 36 women and 67 men, were included in the analysis from the end of 2021 to the end of 2023 at the Bundeswehr Central Hospital in Koblenz. For all patients, ITI was indicated due to refractory oral cortisone use or a contraindication for this and this option was offered to the patients as a possibility. The study was approved by the ethics committee and was conducted in accordance with the ethical guidelines for medical research involving human subjects set out in the Declaration of Helsinki.

The data included in the analysis were all taken from the standardised examination of the patients. No additional data beyond the routine standard was collected for the study.

The patient population consisted of patients who were referred to our clinic for further treatment by ENT colleagues in private practice following refractory oral cortisone treatment. Before each ITI, audiometry was carried out by audiometrists and an ENT examination was performed. This was carried out before each new injection to visualise the progression. Where possible, the injections were given on Monday, Wednesday and Friday. The termination of the injections was not subject to a precise definition, but rather the subjective complaint and the hearing improvement already achieved from the initial situation were weighed against the possible complications of continuing the injections beyond the 10th injection and the patient’s wishes were taken into account.

Injections prepared by the pharmacy with dexamethasone in hyaluronic acid solution (8 mg/ml) are applied; the total of 0.5 ml contains 0.4 ml of Dexa 100 mg Inject and 0.1 ml of Hylo-Comod eye drops. This preparation is prepared under aseptic conditions. The solution is then filled into a syringe (Fig. 2). When stored in a cool place between +2°C and +8°C, these syringes are stable for up to 3 months. This information is based on the results of microbiological and chemical tests.

**Picture 1.**
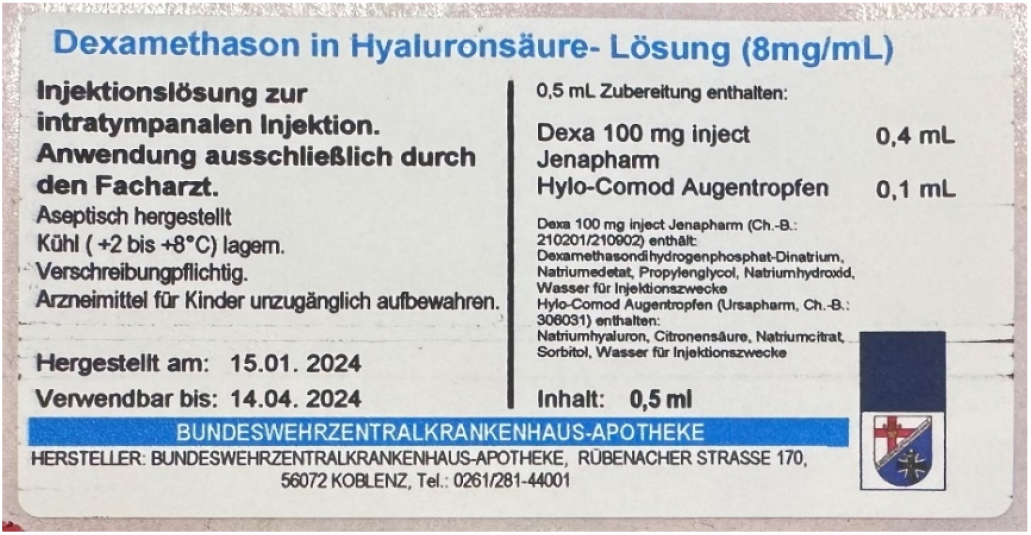
Information on the injection solution

**Picture 2.**
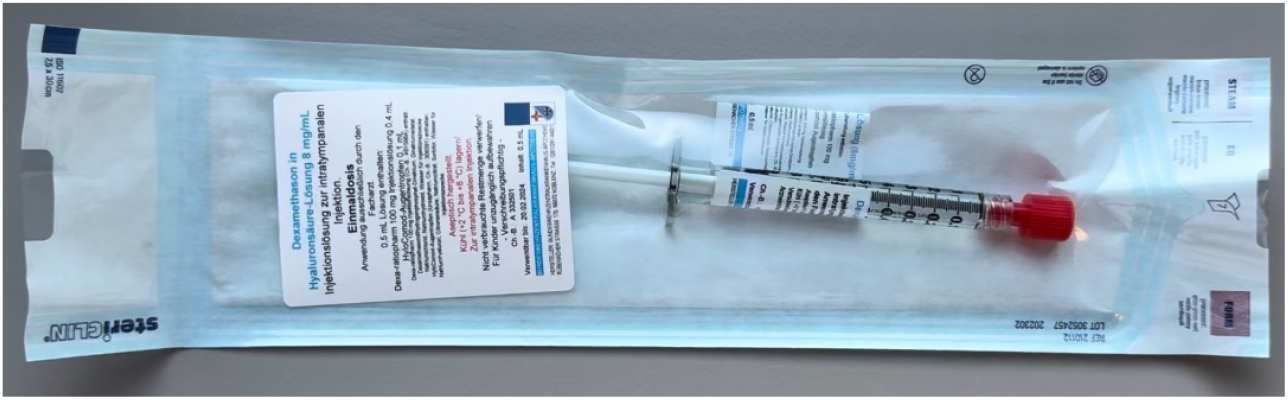
Preparation of pre-filled syringes by the pharmacy for storage in the refrigerator

### 2.2 Indications

The indication was determined in accordance with the S1 guideline on high-dose systemic glucocorticoid therapy and the ITI for the treatment of sudden hearing loss (5). Here, intratympanic cortisone administration is indicated both as primary therapy and as second-line therapy, in the sense of rescue treatment, if the symptoms have not improved sufficiently with first-line therapy, oral cortisone intake, or as first-line therapy as an alternative to oral intake in the presence of contraindications (2). ITI is not recommended for acute isolated tinnitus. In one case in this study, ITI was used contrary to the recommendation due to pronounced distress and patient request.

### 2.3 Application

After microscopic cleaning of the ear canal, the eardrum is visualised under the microscope. The application of lidocaine/prilocaine (25mg/g + 25mg/g) ointment, which was previously filled into a syringe, is now applied to the eardrum and into the ear canal under visualisation using an ear canal aspirator. During this time, the prefabricated syringe, which has been stored in the refrigerator up to this point, is given to the patient to warm up. After a waiting time of 5 minutes, the ointment is sucked out of the ear canal and away from the eardrum. It can already be tested whether the eardrum is anaesthetised. The fluid is then injected into the tympanic cavity again under microscopic control using a 0.60 × 80mm 23G x3 1/8 cannula until a visible mirror forms behind the eardrum and the tympanic cavity appears to be filled. The head is then positioned on the contralateral side for 5 minutes to prevent immediate leakage.

## 3. Results

Of a total of 103 patients who received an ITI, the age meridian was 56 years with a range between 20 and 82 years of age. The right ear was significantly more frequently affected than the left ear. The incidence in males was almost twice as high as in females. 78 patients with sudden deafness were treated, 74 of whom had tinnitus as a concomitant symptom and 10 of whom had concomitant vertigo.

In most patients, ITI was administered as an additive measure when taking an oral cortisone regimen. However, ITI was also given as a second-line measure as rescue therapy in almost the same patient cohort. On average, 8 injections were administered at 3-day intervals.

As a complication, 3 out of 103 patients had a perforation of the eardrum, which had to be surgically closed by means of tympanoplasty. One patient experienced taste disturbances and 7 patients experienced transient symptoms of dizziness. Hearing deterioration occurred in 3 patients, so that ITI treatment was discontinued in these patients.

|  |  |  |
| --- | --- | --- |
|  |  | **n=103** |
| **Age in years** | Median | 56 |
| **Affected page** | Right | 56 |
|  | Links | 47 |
| **Gender** | Male | 67 |
|  | Female | 36 |
| **Complaints or diagnoses** | Tinnitus | 74 |
|  | Barotrauma | 1 |
|  | Blast trauma | 5 |
|  | Hearing impairment or sudden deafness? | 78 |
|  | IED (Improvised Explosive Device) | 2 |
|  | Dizziness | 10 |
| **First-line therapy (with oral cortisone intake)** |  | n=55 |
|  | Start after d (median) | 11 |
|  | Number of injections (median) | 5 |
|  | max. hearing loss (dB) | 110 |
|  | Median hearing improvement (dB) | 10 |
|  | Hearing improvement in mean value (dB) | 20,8 |
| **Second-line therapy (without oral cortisone intake)** |  | n=48 |
|  | Start after d (median) | 7 |
|  | Number of injections (median) | 8 |
|  | max. hearing loss (dB) | 110 |
|  | Median hearing improvement (dB) | 10 |
|  | Hearing improvement in mean value (dB) | 16,9 |
| **Virus serology neurotropic viruses** | Conspicuous | 8 |
| **Complications** | Persistent eardrum perforations | 3 |
|  | Flavour disorders | 1 |
|  | Hearing deterioration | 3 |
|  | Dizziness | 7 |

## 4. Discussion

### 4.1. Indication and duration of treatment

Whether ITI, which acts directly on the cochlea, has a benefit over systemically effective oral cortisone, and whether it can be considered as a primary therapy or whether it offers an additional benefit in patients with inadequate regression of symptoms after systemic cortisone administration, has been widely debated in recent years.

The intraympanic application of medication was already described over 60 years ago (7) and has been used since the treatment of hydropic inner ear diseases (Menièré’s disease) in the mid-1990s as part of intratympanic gentamycin injections (8). The main advantages are the reduction of systemic side effects and the achievement of a higher active level in the inner ear (5).

In 2022, Plontke et al. analysed 30 international studies as part of a Cochrane review to determine the extent to which ITI is beneficial as a therapeutic measure in the treatment of sudden hearing loss. A total of 2133 patient data were analysed. The analyses revealed that ITI as a primary therapy does not show any benefit in terms of hearing improvement in contrast to systemic cortisone administration and that the side effects of intratympanic administration such as pain and dizziness during the injection and resulting eardrum perforations predominate. Combination therapy also showed little or no additional benefit compared to systemic cortisone alone. However, there is not a high level of evidence for this statement, meaning that further studies are required. With regard to ITI in the case of insufficient regression of symptoms after systemic administration (secondary therapy), an improvement in hearing was shown in contrast to placebo or “doing nothing”, which can be assessed as significant (9).

In the HODOKORT study, it was shown that a lower dose of systemic cortisone compared to the recommended dose of the current S1 guideline does not represent a disadvantage. Whether ITI as a secondary therapy in accordance with the recommended dose of the HODOKORT study continues to provide a benefit remains unclear and needs to be investigated.

With reference to the study cited here, the findings from the Cochrane review can be supported. The 103 patient data analysed here also show that the hearing ability of patients who did not take oral cortisone and received only intratympanic treatment showed a lower mean improvement than patients who received both treatment options.

### 4.2. Pharmacokinetics

Compared to systemic administration, higher intracochlear drug levels can be achieved by bypassing the blood-labyrinth barrier during local cortisone administration. After intratympanic administration of the substance, intracochlear distribution occurs by diffusion at the round window membrane. A basoapical concentration gradient occurs in the inner ear (14). Only a small proportion of the glucocorticoid enters the bloodstream. Therefore, systemic side effects are not to be expected. This is particularly advantageous for patients with relevant concomitant diseases, such as diabetes mellitus and hypertension.

### 4.3. Application

Various forms of application with different glucocorticoids have already been discussed. Single or repeated injections have also been discussed. It has not yet been possible to determine how many injections should be administered and when. Whether the application should be given with or without local anaesthesia as an injection or via an inserted tympanostomy tube has also not been clarified. Inspection of the round window niche has also been discussed, but has not yet been conclusively decided to be appropriate. Continuous administration, for example via special pump systems or biodegradable polymers, has been described (14).

As early as 1998, Lange et al. described the change in their own application methods (10). From years of experience with the insertion of small calibre transmeatal tubes or eardrum tubes, as used for the treatment of mucoserotympanum. Transtympanic injection under local anaesthesia is technically simple, safe and quick (11,12).

In 2024, Hoch et al. presented a treatment concept that could be easily implemented in routine clinical practice with three ITIs of dexamethasone 4 mg/ml via direct puncture of the eardrum under local anaesthesia. After approx. 2-4 weeks after the end of unsuccessful systemic high-dose cortisone therapy, the patient is counselled and informed about possible risks. The injections are carried out on 3 consecutive days in an outpatient setting. However, due to the risk of dizziness, the patient should not drive independently on the day in question (13).

Whether and what influence surface anaesthesia could have has not been clarified. The filling of the auditory canal with lidocaine spray, application of ointment to the eardrum and the auditory canal or the insertion of gelatine sponges soaked in local anaesthetic are known (13).

The longer retention of the injected solution by direct injection using a cannula than via a tympanostomy tube can be assumed, but has not been proven. A wide tympanic tube or active swallowing can also cause the premature closure of the injected solution through the auditory tube.

## 5. Conclusion

The results of the study listed here in relation to existing studies show that ITI is beneficial as a supportive therapy after insufficient hearing improvement with systemic cortisone administration. As the sole therapeutic approach for patients who have no contraindication to systemic therapy, it is preferable to ITI.

The advantage of ITI in contrast to high-dose systemic administration is the lack of side effects and the continuous connection of the patient to the doctor. However, the results of the HODOKORT study are likely to lead to a new recommendation regarding the dose of glucocorticoid. Whether the new dose will affect the efficacy of ITI as a possible secondary therapy remains unclear and requires further investigation. The different treatment strategies that currently still exist with regard to indication, implementation, form of application, treatment frequency and duration as well as the substance should be standardised in order to be able to make subsequent statements regarding success.

The retrospective data analysis was evaluated and approved by the ethics committee of the Mainz Medical Association under the number 2024-17577-retrospective.

## Data Availability

There are no limits to publish

## Conflicts of interest

The Author declares that there are no conflicts of interest.

They assure that they have observed and complied with the data protection regulations and the requirements of the Declaration of Helsinki.

